# Cardioprotective effect of the low energy extracorporeal shock wave on the doxorubicin-induced cardiomyopathy in breast cancer patients

**DOI:** 10.1101/2023.06.02.23290908

**Authors:** Shinjeong Song, Joohyun Woo, HyunGoo Kim, Jun Woo Lee, Woosung Lim, Byung-In Moon, Kihwan Kwon

## Abstract

**Background:** Doxorubicin is a highly effective anti-cancer drug but causes left ventricular (LV) dysfunction, and also induces late-onset cardiomyopathy. Extensive research has been done and is being done to discover preventive treatments. However, an effective and clinically applicable preventive treatment is yet to be discovered. Cardiac Extracorporeal shock waves therapy (C-ESWT) has been suggested to treat inflammatory and ischemic diseases and protect cardiomyocytes from doxorubicin-induced cardiomyopathy. The aim of this study is to assess the safety and efficacy of C-ESWT in treating patients with doxorubicin.

**Methods:** A single-center, randomized, prospective controlled study to evaluate the prevention and safety of doxorubicin-induced cardiomyopathy using C-ESWT was performed. We enrolled 64 breast cancer patients. C-ESWT group 33 patients were treated with our cardiac shock wave therapy (200 shots/spot at 0.09mJ/mm2 for 20 spots, 3 times a week every two-chemotherapy cycles). Efficacy endpoints were LVGLS by 2D speckle tracking echocardiography using Tomtec software and CTRCD. Echocardiography should be performed on the baseline line and every 4 cycles of chemotherapy, followed by a follow-up 3,6 months after chemotherapy to compare the incidence of cardiomyopathy of subclinical LV dysfunction due to chemotherapy between the two groups. CTRCD was defined as a reduction in LV ejection fraction (EF) from a baseline greater than 10% to less than 55% and subclinical CTRCD was defined as a GLS reduction of ≥ 15% compared to the baseline value.

**Results:** Participants averaged 50±9 years in age, 100% female. In the results of follow-up 6 months after the end of chemotherapy, there was a significant difference in delta LVGLS between the ESW group and the control group (EF; -0.2±7.7% vs. -3.7±14.2 p-value; 0.076, GLS; -1.2±9.3% vs. -11.2±9.5% p-value; <0.001). In the control group, 2 patients developed CTRCD. A total of 27% (14 patients) of patients developed subclinical LV dysfunction (Control group; 11 vs. ESW group; 3). ESW therapy was performed safely without any serious adverse events.

**Conclusion:** In this prospective study, C-ESWT established efficacy in preventing CRTCD as well as chemotherapy-induced cardiomyopathy using doxorubicin chemotherapy and the safety of C-ESWT use in breast cancer patients.

## Introduction

Doxorubicin is a well-known and widely used anthracycline class therapeutic agent for the treatment of breast cancer, blood cancer, and other types of cancer. Studies using neurohormonal antagonists drugs such as Angiotensin-converting enzyme (ACE) inhibitors or Angiotensin receptor blockers (ARB), beta-blockers, and others have been successful or unsuccessful in preventing doxorubicin-induced cardiomyopathy. Dexrazoxane, which was designated as an orphan drug by the FDA in 2014, is currently the only drug approved for the prevention of anthracycline-induced cardiotoxicity in children and adolescents aged 0-16 years. [1,2]

Currently, guidelines recommend the use of dexrazoxane when using Doxorubicin in children or when an adult has accumulated a certain threshold dose of Doxorubicin. While dexrazoxane is successful in suppressing anthracycline cardiotoxicity, it does not offer complete cardioprotection, since anthracyclines have several possible cardiotoxic mechanisms, and dexrazoxane only targets some of them. [3-5] The cardiotoxicity associated with anthracycline use can range from subclinical cardiomyopathy to heart failure (HF) and even cardiac death. HF may occur within the first week of anthracycline treatment or it may take decades to develop. [6] However, most cases occur within the first year after treatment. [7]

Doxorubicin, an anthracycline used for clinical purposes, has been shown to induce oxidative stress and apoptosis of cardiomyocytes, limiting its long-term use due to cardiotoxicity. [8,9] This is supported by several studies which have demonstrated that doxorubicin increases the risk of cardiotoxicity with cardiac symptoms similar to those of dilated cardiomyopathy. [10-12]. Survivin, a member of the inhibitor of the apoptosis protein family, has been found to regulate cellular apoptosis and tumor progression in various cell types. [13-15] As doxorubicin treatment induces cellular apoptosis in cardiomyocytes, survivin has been identified as a suitable therapeutic target for patients with DOX-induced cardiomyopathy. Therefore, upregulation of endogenous survivin levels may be a more reasonable potential therapy for doxorubicin -induced cardiomyopathy.

Extracorporeal shock wave (ESW) therapy has been used as a first-line treatment for stone diseases due to its high energy. [16,17]. Low-energy ESW has been shown to have protective effects in various diseases associated with bones, tendons, and the musculoskeletal system, and to promote angiogenesis and improve cardiac injury after acute myocardial infarction. [18] Previous studies have shown that low-energy ESW promotes angiogenic gene expression and activates the PI3K/Akt signaling pathway, which is involved in survivin expression.

[13,19,20,21] Based on these findings, subsequent research has proposed ESW as a new specific and safe therapy against acute doxorubicin-induced cardiomyopathy, protecting cardiomyocytes by upregulating survivin in an in vivo model. [22]

Therefore, this study aims to investigate whether Cardiac Extracorporeal Shock Wave Therapy (C-ESWT) can prevent Cancer Therapy-Related Cardiac Dysfunction (CTRCD), especially doxorubicin-induced cardiomyopathy, in patients receiving doxorubicin chemotherapy.

## Methods

Breast cancer patients receiving anthracycline-based chemotherapy were enrolled at Ewha Woman’s Mokdong Hospital between June 2021 and December 2022, with the aim of recruiting at least 72 participants. This study was approved by the institutional review board of Ewha Woman’s Mokdong Hospital (2020-12-044). All participants provided written informed consent before inclusion in the study. The study was registered at https://www.clinicaltrials.gov (NCT05584163) The data that support the findings of this study are available from the corresponding author on reasonable request. Eligible participants had a normal cardiac function, as determined by screening echocardiography, and provided voluntary consent to participate. Participants were randomly assigned to either the treatment group or the control group. Inclusion criteria were patients aged 19 or older, with breast cancer scheduled to receive at least 3 cycles of anthracycline-based chemotherapy, and normal cardiac function. Exclusion criteria were patients with structural heart disease, intracardiac device presence, antiarrhythmic drugs that may affect the QT interval, and recent defibrillation due to AF or previous PCI or coronary artery bypass surgery.

A total of 64 patients were randomly divided into the ESWT group (n=33) and control group (n=31), with 8 patients excluded due to declining to participate or inconvenient transportation. Patients in the ESWT group received ESWT and standard chemotherapy per guidelines, while those in the control group received standard chemotherapy containing doxorubicin.

### C-ESWT protocol

C-ESWT was administered three times a week, on the first, third, and fifth days of each course, with a 6-week interval between two courses. Each patient received at least six sessions of C-ESWT. The energy and the total number of ESWT therapy were set to 200 shots/spot at 0.09mJ/mm^2^ for 40 spots per session. This protocol has been proven safe in previous studies targeting patients with coronary artery disease and refractory angina. [23,24] During the C-ESWT procedure, the location of the left ventricular myocardium was confirmed using echocardiography, and C-ESWT was performed. Additionally, an EKG was connected to monitor arrhythmia. At baseline and follow-up, peripheral venous blood samples were collected and analyzed for myocardial markers including creatine kinase (CK), creatine kinase phosphate-isozyme (CK-MB), and Troponin T (TnT). Additionally, the Complete Blood Count (CBC) and hepatorenal function indexes alanine aminotransferase (ALT), aspartate aminotransferase (AST), and lactic dehydrogenase (LDH) serum creatinine (Cr) were also measured.

### Echocardiography

Patients underwent echocardiography at baseline prior to starting anthracycline therapy, and again at 3 and 6 months after completing chemotherapy, which included doxorubicin. The echocardiograms were obtained using GE Ultrasound System and Philips Ultrasound Machines and included 3 to 5 cardiac cycles of 4-, 3-, and 2-chamber apical views. The images were digitally stored in raw format for later analysis. LVGLS was analyzed using a semiautomated speckle tracking technique (TOMTEC Imaging system GmbH Freisinger Strasse9, 85716 Unterschleissheim Germany) with a model of the entire left ventricle, including the 3 apical views. Inadequately tracked segments were excluded from the analysis. Two different observers analyzed the datasets and measured the left ventricular global longitudinal strain (LVGLS) using TOMTEC. Each observer analyzed the dataset twice, with the second review taking place 3 months after the first. The observers were blinded to each other during the analysis, and the second measurement of each observer was used to evaluate intraobserver reproducibility.

### Outcome

The primary outcome was the difference in LVGLS between baseline and the 3-month and 6-month follow-up in the ESWT group compared to the control group. The secondary outcome was the difference in the rate of CTRCD between the two groups. In this study, CTRCD was defined as a reduction in LVEF of >10 absolute percentage points to a value <50% as measured by echocardiography at any follow-up time point [25]. These patients were defined as having definite CTRCD. Probable CTRCD was defined as a reduction in LVEF of >10 absolute percentage points to a value ≥50% accompanied by a relative reduction in LVGLS of >15%. Possible CTRCD was defined as a reduction in LVEF of <10 absolute percentage points to a value <50% or a relative reduction in GLS of ≥15% from baseline. This study defined patients meeting these criteria as having possible CTRCD. [25]

### Statistical Analysis

The statistical analysis was carried out using SPSS version 23.0 (SPSS Inc., USA). Continuous variables with a normal distribution were reported as mean ± standard deviation and analyzed using paired t-tests for baseline and follow-up comparison. Categorical variables were presented as frequency (n) or ratio and analyzed using the chi-square test. Rank data were analyzed using a non-parametric rank sum test. Intraobserver and interobserver reliabilities were measured using the intraclass correlation coefficient and the Bland-Altman analysis. A p-value of less than 0.05 (two-tailed) was considered statistically significant.

## Results

### Patient Characteristics

Out of the 72 patients who were randomized, 3 (7%) did not receive follow-up (as shown in Figure 1). Five patients withdrew their consent or did not come back for imaging follow-up after the baseline study. The final analysis included 64 patients (33 in the ESWT treatment group and 31 in the control group). A comparison of the baseline clinical and cardiac imaging characteristics of the included patients between the two groups is presented in Table 1. All patients were women, with an average age of 50 ± 9 years (range 29 to 72 years). Common risk factors for heart failure were: 11 (17.2%) had hypertension, 4 (6.3%) had diabetes mellitus, 5 (7.8%) had dyslipidemia, and none were current or ex-smokers. There was no significant difference in baseline EF or LVGLS between the two groups. All patients had breast cancer and received anthracycline-based chemotherapy. The median doxorubicin equivalent dose was 241 mg/m2 (interquartile range [IQR]: 238 to 245 mg/m2), and there was no significant difference between the C-ESWT and control groups (C-ESWT group: 241 mg/m2 [IQR: 237 to 244 mg/m2]; control group: 241 mg/m2 [IQR: 238 to 245 mg/m2]; p = 0.14).

**Table 1.** Patient Characteristics.

|  | ESWT group<br>(N=33) | Control group<br>(N=31) | P value |
| --- | --- | --- | --- |
| Demographics |  |  |  |
| Age, y | 51±7 | 50±11 | 0.672 |
| Female | 34 (100) | 32 (100) | - |
| Risk factors |  |  |  |
| Diabetes | 4 (11.8) | 1 (3.1) | 0.190 |
| Hypertension | 7 (20.6) | 5 (15.6) | 0.608 |
| Dyslipidemia | 4 (11.8) | 2 (6.3) | 0.440 |
| Smoking | 0 | 0 | - |
| BMI, kg/m<sup>2</sup> | 23.7±5.0 | 22.7±4.0 | 0.363 |
| Previous cardiovascular disease | 0 | 0 | - |
| Baseline medical therapy |  |  |  |
| Beta-blocker | 0 | 1 (3.1) | 0.306 |
| ACEI or ARB | 6 (17.6) | 2 (6.3) | 0.161 |
| Statin | 4 (11.8) | 2 (6.3) | 0.440 |
| Observations |  |  |  |
| Systolic blood pressure, mmHg | 121±15 | 119±15 | 0.539 |
| Diastolic blood pressure, mmHg | 73±11 | 73±10 | 0.974 |
| Heart rate, beats/min | 73±11 | 73±14 | 0.941 |
| Chemotherapy |  |  | 0.112 |
| Neoadjuvant | 5 (14.7) | 10 (31.3) |  |
| Adjuvant | 29 (85.3) | 22 (68.8) |  |
| Additional therapy after Doxorubicin |  |  | 0.647 |
| Paclitaxel | 30 (88.2) | 28 (87.5) |  |
| Docetaxel | 3 (8.8) | 1 (3.1) |  |
| Trastuzumab | 0 | 2 (6.3) |  |
| Letrozole | 1 (2.9) | 1 (3.1) |  |
| Cumulative doxorubicin dose, mg/m<sup>2</sup> | 240 ± 6 | 243 ± 10 | 0.133 |
| EF |  |  |  |
| Baseline, % | 65±5 | 66±4 | 0.366 |
| GLS |  |  |  |
| Baseline, % | -20.9 ± 2.6 | -20.7 ± 2.2 | 0.751 |
| Baseline < - 18% | 31 (90.9) | 29 (90.6) |  |
| Baseline -16% ~ -18% | 3 (9.1) | 3 (9.4) |  |
| Baseline >-16% | 0 | 0 |  |
| Laboratory |  |  |  |
| Baseline CK | 54 ± 25 | 68 ± 33 | 0.366 |
| Baseline CK-MB | 1.3 ± 1.2 | 1.0 ± 0.4 | 0.610 |
| Baseline Troponin T | 0.01 ± 0.011 | 0.02 ± 0.014 | 0.384 |

**Figure 1.**
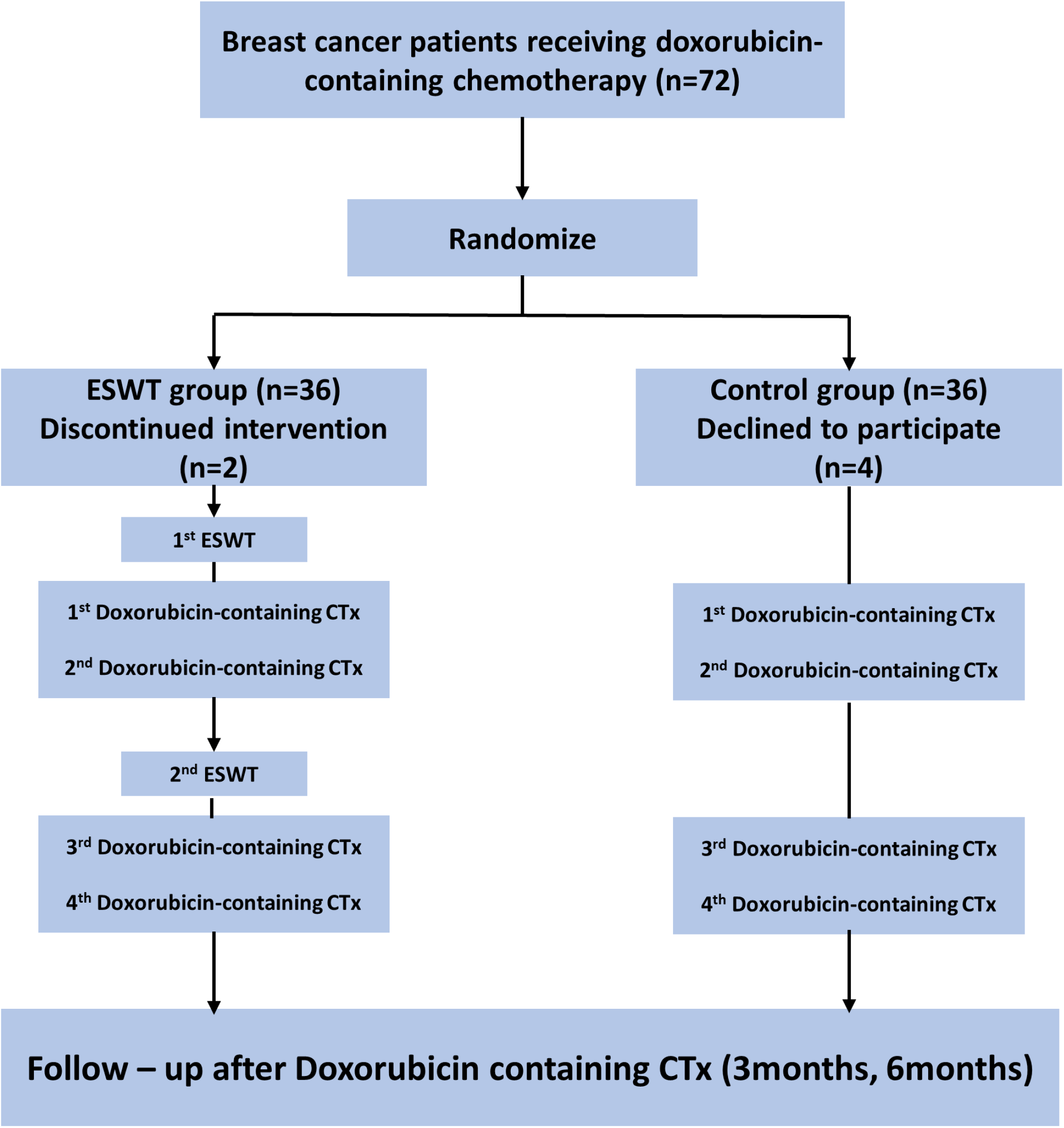
Prevention of CTRCD (doxorubicin-induced cardiomyopathy) protocol in the C-ESWT group and control group CTRCD; Cancer Therapy-Related Cardiac Dysfunction, C- ESWT; Cardiac Extracorporeal shock wave therapy

### Primary outcome – LVGLS

There was no significant difference in baseline LVGLS between the two groups (C-ESWT -20.9% ± 2.6 vs. bcontrol group -20.7% ± 2.2, p-value 0.751). However, at 3 months after anthracycline-based chemotherapy, including doxorubicin, the GLS values were -20.0% ± 2.8 and -17.9% ± 2.6 for the C-ESWT and control groups, respectively. These values showed a statistically significant difference in the change from baseline, with -4.1% vs. -13.5%, respectively. A similar trend was observed at 6 months, where the GLS values also showed statistically significant differences between the two groups and from baseline (Table 2, Figure 2). However, there was no significant difference in LVEF, LVEDVi, and LVESVi between the two groups at 3 and 6 months (Table 2).

**Table 2.** EF and LVGLS measures throughout Doxorubicin therapy.

|  | ESWT group<br>(N=33) | Control group<br>(N=31) | P value |
| --- | --- | --- | --- |
| LVEF, % |  |  |  |
| Baseline | 64.8 ± 4.9 | 66.3 ± 4.3 | 0.213 |
| Post - 3months | 64.7 ± 4.4 | 62.5 ± 7.4 | 0.150 |
| Change from baseline (%) | 0.0 ± 5.7 | -5.7 ± 10.1 | 0.008 |
| Post - 6months | 64.7 ± 4.4 | 63.5 ± 5.9 | 0.413 |
| Change from baseline (%) | -0.2 ± 7.7 | -3.7 ± 7.3 | 0.076 |
| LVEDVi, mL/m2 |  |  |  |
| Baseline | 42.5 ± 8.7 | 44.7 ± 10.9 | 0.371 |
| Post - 3months | 43.1 ± 9.2 | 43.3 ± 10.7 | 0.942 |
| Change from baseline (%) | 2.9 ± 18.9 | -0.9 ± 21.8 | 0.458 |
| Post - 6months | 43.9 ± 10.2 | 41.4 ± 9.3 | 0.359 |
| Change from baseline (%) | 5.3 ± 21.5 | -5.7 ± 20.4 | 0.067 |
| LVESVi, mL/m2 |  |  |  |
| Baseline | 17.6 ± 5.3 | 19.2 ± 6.2 | 0.269 |
| Post - 3months | 18.6 ± 4.8 | 21.6 ± 6.4 | 0.036 |
| Change from baseline (%) | 11.4 ± 34.5 | 16.9 ± 30.2 | 0.501 |
| Post - 6months | 18.4 ± 5.3 | 20.5 ± 5.3 | 0.161 |
| Change from baseline (%) | 13.0 ± 41.4 | 10.5 ± 25.5 | 0.795 |
| GLS, % |  |  |  |
| Baseline | -20.9 ± 2.6 | -20.7 ± 2.2 | 0.751 |
| Post - 3months | -20.0 ± 2.8 | -17.9 ± 2.6 | 0.002* |
| Change from baseline (%) | -4.1 ± 8.6 | -13.5 ± 9.3 | <0.001* |
| Post - 6months | -20.5 ± 1.9 | -18.4 ± 2.5 | <0.001* |
| Change from baseline (%) | -1.2 ± 9.3 | -11.2 ± 9.5 | <0.001* |

\*p value <0.05
LVEF; Left ventricular ejection fraction, LVGLS; Left Ventricular Longitudinal Global Strain

**Figure 2.**
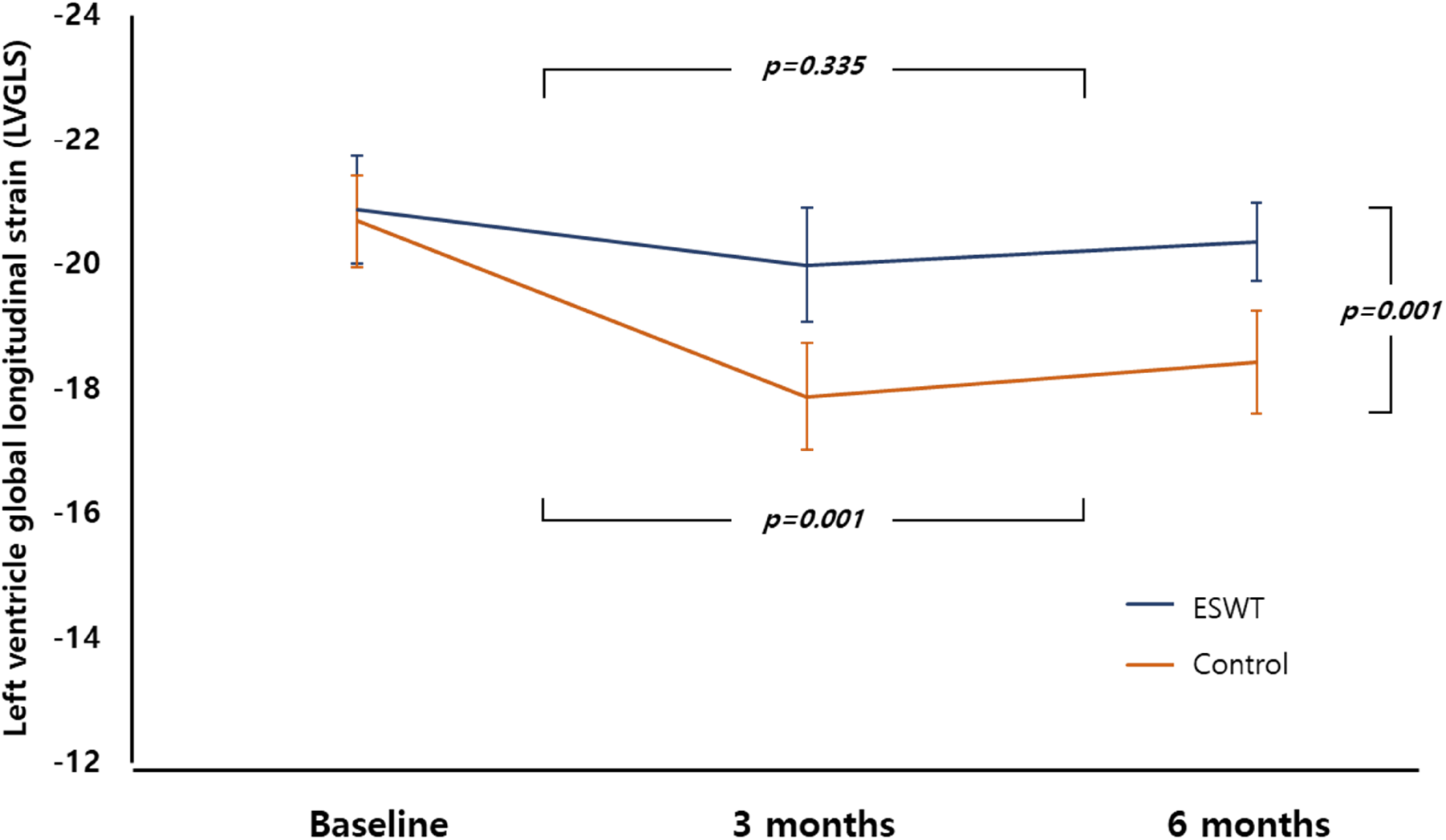
Trend of LVGLS at baseline, 3months, 6months between the two groups LVGLS; Left Ventricular Longitudinal Global Strain

### Secondary outcome – CTRCD

There were no significant differences in LVEF or LVGLS between the two groups at baseline, as shown in Table 1.Cardiotoxicity occurring at any time during the study was observed only in two patients in the control group.

After completion of anthracycline-based chemotherapy, probable CTRCD was not observed in either group. But possible CTRCD occurred in two patients in the control group at 6 months and in 11 patients in the ESWT group, demonstrating statistical significance between the two groups. The occurrence of any type of CTRCD during follow-up was significantly higher in the control group than in the C-ESWT group (13 events vs. 2 events; p-value = 0.001) as shown in Table 3.

**Table 3.** Cancer Therapy-Related Cardiac Dysfunction (CTRCD) between ESWT and Control group.

|  | ESWT group<br>(N=33) | Control group<br>(N=31) | P value |
| --- | --- | --- | --- |
| Total | 2 | 13 | 0.001 |
| Cardiotoxicity | 0 | 2 | 0.143 |
| Probable subclinical cardiotoxicity | 0 | 0 | - |
| Possible subclinical cardiotoxicity | 2 | 11 | 0.003 |

CTRCD; Cancer Therapy-Related Cardiac Dysfunction, C- ESWT; Cardiac Extracorporeal shock wave therapy

### Safety

All patients tolerated ESWT well without any major complications. Only 3 patients (9%) reported pain around the C-ESWT site. However, no bruises or skin lesions were observed at the site where extracorporeal shock wave therapy was performed. Throughout the trial, there were no clinically significant increases in BNP, CK, CK-MB, or troponin I level in individual patients. The mean BNP levels at screening, 3-month, and 6-month follow-up were 172.8±286.9, 147.9±303.8, and 173.7±299.1 pg/mL, respectively, and there were no significant changes in ECGs from baseline to follow-up.

## Discussion

This pilot study suggests that C-ESWT therapy is a safe, non-invasive, and effective prevention method for doxorubicin-induced cardiomyopathy, for patients with breast cancer, for which no established preventive drug or method is currently available. In the C-ESWT group, there was a significantly lower incidence of CTRCD, and no serious adverse events related to extracorporeal shock wave therapy were observed.

Doxorubicin is known for inducing oxidative stress and cardiomyocyte apoptosis, resulting in cardiotoxicity. A previous article proposed survivin, an inhibitor of apoptosis protein, as a promising target for treating DOX-induced cardiomyopathy since it regulates cellular apoptosis. Using ESWT promotes an increase in survivin, and its cardioprotective effect has already been demonstrated in in vivo studies. This study is significant in confirming the safety and myocardial protective effect of ESWT through LVGLS analysis in humans.

The previous studies that used ESWT on the heart mostly aimed to improve symptoms or left ventricular systolic function (LVEF) in refractory angina patients. [23,24,26] However, this study is the first to investigate whether C-ESWT can prevent doxorubicin-induced cardiomyopathy in a homogeneous group of female breast cancer patients. Doxorubicin is an effective anticancer drug for breast cancer, sarcoma, and hematologic malignancies, but its use is limited due to its toxicity, particularly cardiac toxicity. Although there are ongoing efforts to investigate the efficacy of neurohormonal antagonists, such as renin-angiotensin-aldosterone inhibitors and beta-blockers, in reducing cardiotoxicity, their routine use has not yet been widely adopted in clinical practice due to inconsistent results. [27-32] Guidelines and the US FDA do not provide indications for using dexrazoxane to prevent cardiac toxicity in adult patients who use doxorubicin at non-high doses or from the outset. Therefore, there is currently no preventive method for the risk of cardiomyopathy that may occur with the use of low-dose doxorubicin.

The possible and probable cardiomyopathy definition criteria presented in recent guidelines utilize LVGLS. LVGLS is known to predict future EF reduction when it significantly decreases before EF reduction in patients undergoing chemotherapy. [33] LVGLS is widely used because it is well-correlated with prognosis in not only CTRCD but also heart failure, valvular disease, and myocardial disease. Although LVGLS does not meet the definition of definite CMP in CTCRD, it has shown a meaningful reduction in LVGLS in probable and possible CMP cases, which may lead to the development of overt heart failure in long-term follow-up. Therefore, ESWT has proven to be a useful tool in preventing possible and probable cardiomyopathy in two groups, and if long-term follow-up results confirm this, it may serve as robust evidence for the prevention of CTRCD using C-ESWT.

In previous studies, doxorubicin-induced cardiotoxicity was known to increase in frequency with a higher cumulative dose. [36] However, even in patients not receiving high-dose doxorubicin chemotherapy, there is a significant incidence of probable cardiomyopathy according to the CTRCD criteria. [34] This indicates the need for preventive measures to avoid cardiotoxicity from the outset, not only in high-dose doxorubicin-based chemotherapy, as recommended by the guidelines that allow the use of dexrazoxane but also in cases of low-dose doxorubicin-based chemotherapy in adult cancer patients.

To ensure safety during shock wave therapy, continuous EKG monitoring was conducted to detect arrhythmias, and there were no new cases of arrhythmia in subsequent follow-up EKG. Furthermore, there were no patients with suspected hemolysis anemia in CBC performed serially during chemotherapy, and no serious adverse events occurred.

### Study strengths and limitations

To our knowledge, our study is the first randomized controlled study to investigate the preventative effects of C-ESWT on CTRCD in patients receiving anthracycline-based cancer therapy. A significant strength of our study is the use of highly standardized entry criteria, resulting in a relatively homogeneous patient population, despite the small sample size. Additionally, we focused on patients with relatively low heart failure risk factors. To overcome the differences in LVGLS between vendors, all images were anonymized and analyzed using the TOMTEC program. In addition, the analysis was performed twice by two investigators, and the strengths of the study include confirming the LVGLS trend at 3 and 6 months after the completion of anticancer therapy. As is well known, our study also demonstrated a high level of agreement in LVGLS measurements both between and within examiners. (Supplementary) Although our data suggest limited adverse events associated with C-ESWT therapy, caution must be exercised in interpreting these results due to the small sample size. Larger multicenter confirmatory studies, including sham-controlled randomized trials, are needed to ascertain the beneficial effect of ESWT in patients receiving anthracycline-based cancer therapy.

However, due to the limited number of participants, the available data indicate that C-ESWT has few adverse events, and the present data must be interpreted with caution to obtain a more accurate understanding of the potential benefits of this technique, larger standardized studies are needed. Therefore, larger multicenter confirmatory studies, which include randomized trials with sham control, are necessary to confirm the beneficial effects of C-ESWT in patients undergoing anthracycline-based cancer therapy. Also, although there was no statistically significant difference between the two groups in definite cardiomyopathy, it only occurred in the control group. The incidence of definite cardiomyopathy that occurred in the control group did not show a significant difference compared to other studies, and the incidence of probable and possible cardiomyopathy was not considered excessively high compared to some previous studies. [35,36] However, this also has the limitation of a small overall sample size. The absence of definite cardiomyopathy in the C-ESWT group can be explained by previous studies, which showed that C-ESWT was effective in treating acute doxorubicin-induced cardiomyopathy. [22]

## Conclusion

In conclusion, this pilot study demonstrates that Cardiac Extracorporeal Shock Wave Therapy is a safe and effective method for preventing CRTCD (especially doxorubicin-induced cardiomyopathy) in breast cancer patients. The safety of C-ESWT therapy was confirmed, as no serious adverse events were observed. These findings have important implications for the prevention of doxorubicin-induced cardiomyopathy and the potential use of C-ESWT therapy as a preventive measure.

## Data Availability

The data that support the findings of this study are available from the corresponding author on reasonable request.

## Abbreviations

LVGLS: Left Ventricular Longitudinal Global Strain
ESWT: Extracorporeal shock wave therapy
C-ESWT: Cardiac Extracorporeal shock wave therapy
CTRCD: Cancer Therapy-Related Cardiac Dysfunction

## Acknowledgments

This work was supported by the Korea Medical Device Development Fund grant funded by the Korea government (the Ministry of Science and ICT, the Ministry of Trade, Industry and Energy, the Ministry of Health & Welfare, the Ministry of Food and Drug Safety) (Project Number: 1711139087, RS-2021-KD000005)

## Reference

1 Armenian SH, Ehrhardt MJ. Optimizing Cardiovascular Care in Children With Acute Myeloid Leukemia to Improve Cancer-Related Outcomes. J Clin Oncol. 2019;37:1–6. doi:10.1200/JCO.18.01421

2 US Food and Drug Administration. Orphan drug designations and approvals https://www.accessdata.fda.gov/scripts/opdlisting/oopd/detailedIndex.cfm?cfgridkey=441314.)

3 Lipshultz SE, Herman EH. Anthracycline cardiotoxicity: the importance of horizontally integrating pre-clinical and clinical research. Cardiovasc Res. 2018;114:205–209. doi:10.1093/cvr/cvx246

4 Renu K, V G A, P B TP, Arunachalam S. Molecular mechanism of doxorubicin-induced cardiomyopathy - An update. Eur J Pharmacol. 2018;818:241–253. doi:10.1016/j.ejphar.2017.10.043

5 Cappetta, D., Rossi, F., Piegari, E., Quaini, F., Berrino, L., Urbanek, K., & De Angelis, A. Doxorubicin targets multiple players: A new view of an old problem. Pharmacol Res. 2018;127:4–14. doi:10.1016/j.phrs.2017.03.016

6 Lipshultz SE, Alvarez JA, Scully RE. Anthracycline associated cardiotoxicity in survivors of childhood cancer. Heart. 2008;94(4):525–533. doi:10.1136/hrt.2007.136093

7 Cardinale D, Colombo A, Bacchiani G, Tedeschi I, Meroni CA, Veglia F, Civelli M, Lamantia G, Colombo N, Curigliano G, Fiorentini C, Cipolla CM. Early detection of anthracycline cardiotoxicity and improvement with heart failure therapy. Circulation. 2015;131:1981–8. doi: 10.1161/CIRCULATIONAHA.114.013777.

8 Bartlett JJ, Trivedi PC, Pulinilkunnil T. Autophagic dysregulation in doxorubicin cardiomyopathy. J Mol Cell Cardiol. 2017;104:1–8. doi:10.1016/j.yjmcc.2017.01.007

9 Li X, Liu M, Sun R, Zeng Y, Chen S, Zhang P. Cardiac complications in cancer treatment - A review. Hellenic J Cardiol. 2017;58:190–193. doi:10.1016/j.hjc.2016.12.003

10 Chatterjee K, Zhang J, Honbo N, Karliner JS. Doxorubicin cardiomyopathy. Cardiology. 2010;115:155–162. doi:10.1159/000265166

11 Kumar S, Marfatia R, Tannenbaum S, Yang C, Avelar E. Doxorubicin-induced cardiomyopathy 17 years after chemotherapy. Tex Heart Inst J. 2012;39:424–427.

12 Sandhu H, Maddock H. Molecular basis of cancer-therapy-induced cardiotoxicity: introducing microRNA biomarkers for early assessment of subclinical myocardial injury. Clin Sci (Lond). 2014;126:377–400. doi:10.1042/CS20120620

13 Lee BS, Kim SH, Oh J, Jin T, Choi EY, Park S, Lee SH, Chung JH, Kang SM. C-reactive protein inhibits survivin expression via Akt/mTOR pathway downregulation by PTEN expression in cardiac myocytes. PLoS One. 2014;9:e98113. doi: 10.1371/journal.pone.0098113.

14 Marusawa H, Matsuzawa S, Welsh K, Zou H, Armstrong R, Tamm I, Reed JC. HBXIP functions as a cofactor of survivin in apoptosis suppression. EMBO J. 2003;22:2729–40. doi: 10.1093/emboj/cdg263.

15 Song Z, Yao X, Wu M. Direct interaction between survivin and Smac/DIABLO is essential for the anti-apoptotic activity of survivin during taxol-induced apoptosis. J Biol Chem. 2003;278:23130–40. doi: 10.1074/jbc.M300957200.

16 Argyropoulos AN, Tolley DA. Optimizing shock wave lithotripsy in the 21st century. Eur Urol. 2007;52:344–52. doi: 10.1016/j.eururo.2007.04.066.

17 McClain PD, Lange JN, Assimos DG. Optimizing shock wave lithotripsy: a comprehensive review. Rev Urol. 2013;15:49–60

18 Schmitz C, Császár NB, Milz S, Schieker M, Maffulli N, Rompe JD, Furia JP. Efficacy and safety of extracorporeal shock wave therapy for orthopedic conditions: a systematic review on studies listed in the PEDro database. Br Med Bull. 2015;116:115–38. doi: 10.1093/bmb/ldv047.

19 Ha CH, Kim S, Chung J, An SH, Kwon K. Extracorporeal shock wave stimulates expression of the angiogenic genes via mechanosensory complex in endothelial cells: mimetic effect of fluid shear stress in endothelial cells. Int J Cardiol. 2013;168:4168–77. doi: 10.1016/j.ijcard.2013.07.112.

20 Yu W, Shen T, Liu B, Wang S, Li J, Dai D, Cai J, He Q. Cardiac shock wave therapy attenuates H9c2 myoblast apoptosis by activating the AKT signal pathway. Cell Physiol Biochem. 2014;33:1293–303. doi: 10.1159/000358697.

21 Lee BS, Oh J, Kang SK, Park S, Lee SH, Choi D, Chung JH, Chung YW, Kang SM. Insulin Protects Cardiac Myocytes from Doxorubicin Toxicity by Sp1-Mediated Transactivation of Survivin. PLoS One. 2015;10:e0135438. doi: 10.1371/journal.pone.0135438.

22 Yoon Lee J, Chung J, Hwa Kim K, Hyun An S, Yi JE, Ae Kwon K, Kwon K. Extracorporeal shock waves protect cardiomyocytes from doxorubicin-induced cardiomyopathy by upregulating survivin via the integrin-ILK-Akt-Sp1/p53 axis. Sci Rep. 2019;9:12149. doi: 10.1038/s41598-019-48470-0.

23 Kikuchi Y, Ito K, Ito Y, Shiroto T, Tsuburaya R, Aizawa K, Hao K, Fukumoto Y, Takahashi J, Takeda M, et al. Double-blind and placebo-controlled study of the effectiveness and safety of extracorporeal cardiac shock wave therapy for severe angina pectoris. Circ J. 2010;74:589–91. doi: 10.1253/circj.cj-09-1028.

24 Cassar A, Prasad M, Rodriguez-Porcel M, Reeder GS, Karia D, DeMaria AN, Lerman A. Safety and efficacy of extracorporeal shock wave myocardial revascularization therapy for refractory angina pectoris. Mayo Clin Proc. 2014;89:346–54. doi: 10.1016/j.mayocp.2013.11.017.

25 Dobson R, Ghosh AK, Ky B, Marwick T, Stout M, Harkness A, Steeds R, Robinson S, Oxborough D, Adlam D, et al.; British Society of Echocardiography (BSE) and theBritish Society of Cardio-Oncology (BCOS). BSE and BCOS Guideline for Transthoracic Echocardiographic Assessment of Adult Cancer Patients Receiving Anthracyclines and/or Trastuzumab. JACC CardioOncol. 2021;3:1–16. doi: 10.1016/j.jaccao.2021.01.011.

26 Graber M, Nägele F, Hirsch J, Pölzl L, Schweiger V, Lechner S, Grimm M, Cooke JP, Gollmann-Tepeköylü C, Holfeld J. Cardiac Shockwave Therapy - A Novel Therapy for Ischemic Cardiomyopathy? Front Cardiovasc Med. 2022;9:875965. doi: 10.3389/fcvm.2022.875965.

27 Kalay N, Basar E, Ozdogru I, Er O, Cetinkaya Y, Dogan A, Inanc T, Oguzhan A, Eryol NK, Topsakal R, et al. Protective effects of carvedilol against anthracycline-induced cardiomyopathy. J Am Coll Cardiol. 2006;48:2258–62. doi: 10.1016/j.jacc.2006.07.052.

28 Bosch X, Rovira M, Sitges M, Domènech A, Ortiz-Pérez JT, de Caralt TM, Morales-Ruiz M, Perea RJ, Monzó M, Esteve J. Enalapril and carvedilol for preventing chemotherapy-induced left ventricular systolic dysfunction in patients with malignant hemopathies: the OVERCOME trial (preventiOn of left Ventricular dysfunction with Enalapril and caRvedilol in patients submitted to intensive ChemOtherapy for the treatment of Malignant hEmopathies). J Am Coll Cardiol. 2013;61:2355–62. doi: 10.1016/j.jacc.2013.02.072.

29 Cardinale D, Colombo A, Sandri MT, Lamantia G, Colombo N, Civelli M, Martinelli G, Veglia F, Fiorentini C, Cipolla CM. Prevention of high-dose chemotherapy-induced cardiotoxicity in high-risk patients by angiotensin-converting enzyme inhibition. Circulation. 2006;114:2474–81. doi: 10.1161/CIRCULATIONAHA.106.635144.

30 Akpek M, Ozdogru I, Sahin O, Inanc M, Dogan A, Yazici C, Berk V, Karaca H, Kalay N, Oguzhan A, et al. Protective effects of spironolactone against anthracycline-induced cardiomyopathy. Eur J Heart Fail. 2015;17:81–9. doi: 10.1002/ejhf.196.

31 Gulati G, Heck SL, Ree AH, Hoffmann P, Schulz-Menger J, Fagerland MW, Gravdehaug B, von Knobelsdorff-Brenkenhoff F, Bratland Å, StorÅs TH, et al. Prevention of cardiac dysfunction during adjuvant breast cancer therapy (PRADA): a 2 × 2 factorial, randomized, placebo-controlled, double-blind clinical trial of candesartan and metoprolol. Eur Heart J. 2016;37:1671–80. doi: 10.1093/eurheartj/ehw022.

32 Georgakopoulos P, Roussou P, Matsakas E, Karavidas A, Anagnostopoulos N, Marinakis T, Galanopoulos A, Georgiakodis F, Zimeras S, Kyriakidis M, et al. Cardioprotective effect of metoprolol and enalapril in doxorubicin-treated lymphoma patients: a prospective, parallel-group, randomized, controlled study with 36-month follow-up. Am J Hematol. 2010;85:894–6. doi: 10.1002/ajh.21840.

33 Thavendiranathan P, Poulin F, Lim KD, Plana JC, Woo A, Marwick TH. Use of myocardial strain imaging by echocardiography for the early detection of cardiotoxicity in patients during and after cancer chemotherapy: a systematic review. J Am Coll Cardiol. 2014;63:2751–68. doi: 10.1016/j.jacc.2014.01.073.

34 Barrett-Lee PJ, Dixon JM, Farrell C, Jones A, Leonard R, Murray N, Palmieri C, Plummer CJ, Stanley A, Verrill MW. Expert opinion on the use of anthracyclines in patients with advanced breast cancer at cardiac risk. Ann Oncol. 2009;20:816–27. doi: 10.1093/annonc/mdn728.

35 Lee SH, Cho I, You SC, Cha MJ, Chang JS, Kim WD, Go KY, Kim DY, Seo J, Shim CY, et al. Cancer Therapy-Related Cardiac Dysfunction in Patients Treated with a Combination of an Immune Checkpoint Inhibitor and Doxorubicin. Cancers (Basel). 2022;14:2320. doi: 10.3390/cancers14092320.

36 Tan TC, Bouras S, Sawaya H, Sebag IA, Cohen V, Picard MH, Passeri J, Kuter I, Scherrer-Crosbie M. Time Trends of Left Ventricular Ejection Fraction and Myocardial Deformation Indices in a Cohort of Women with Breast Cancer Treated with Anthracyclines, Taxanes, and Trastuzumab. J Am Soc Echocardiogr. 2015 ;28:509–14. doi: 10.1016/j.echo.2015.02.001.

